# Performance validation of the PRADA digital care system in a cohort-controlled study demonstrates improved end-of-life registration associated with reduced adverse outcomes

**DOI:** 10.64898/2026.01.01.26343327

**Authors:** Baldev M Singh, The Wolverhampton Digital Heath Primary Care Research Network, Amy Thompson, Emily Heyting, Vijay Klaire, Jonathan Lampitt, Emma Parry

## Abstract

Rising urgent care use, avoidable hospital deaths, and missed end-of-life (EOL) registration reflect poor care at this critical phase of life. These are known to be avoidable by earlier identification and EOL registration for effective care planning. Current identification tools are generally non-digital. PRADA, a digital system, uses prediction methodologies for early identification of individuals, escalated to clinicians in active decision support, to promote clinical assessment, the completion of EOL process, EOL registration and to improve outcomes. Our objective was to assess its effectiveness.

In 12 participating general practices, baseline EOL metrics were assessed retrospectively over 2 years (n=64,887, alive 63,344 (97.6%), died 1,543 (2.4%)). Following implementation of the PRADA system outcomes were determined prospectively over 1 -year (n= 66,321 (alive 65,496 (98.7%), died 831 (1.3%)).

EOL registration in those that died, versus non-registration, was associated with lower A/E attendances, emergency admissions, hospital bed day occupancy (all p<0.001) and markedly reduced deaths in hospital (18.5% vs 51.0%, p<0.001). Implementation of PRADA improved registration rates (41.8% vs 25.8%, p<0.001). This shift, for the final year cohort (n=831), identified an additional 133 deaths, avoided 24 deaths-in-hospital, saved 386 bed days, with an estimated direct cost avoidance of £192,789. EOL registration increased in participating practices by 17.6% ±13.7% (range -1.4 to +52.6). However, A/E attendance (74.4%) and urgent admission rates (70.6%) remained high in those registered, each indicating scope for further improvements and efficiency gains over time.

EOL registration rates were low at baseline, missed registration was associated with adverse outcomes, the use of PRADA improved EOL registration by 62% (41.8% vs 25.8%). PRADA use at large scale, in real time direct care demonstrates strong utility, benefit, and performance validity.

**Author Summary:** Existing screening tools for identifying patients with potential palliative care needs are generally manual and paper based, developed in small or disease specific cohorts, or in hospital settings. Systematic reviews show only moderate performance in accurate mortality prediction, with weak evidence for effective deployment in real time care and little validity testing.

In this study, end-of-life registration in those that died was associated with reduced urgent care events, hospital bed days occupied and deaths in hospital. Large-scale deployment of PRADA, a real-time end-of-life digital health care system, over 1-year general practitioners was associated with a 62% increase in registration rates and reductions of adverse events. The study highlights between practice variation, proposing mechanisms for its mitigation with newly defined benchmarks. PRADA thus has high utility, strong clinical benefit and robust performance validity.

The UK NHS 10-year plan has called for better care at the end-of-life and calls for digital solutions. Current tools do not have digital capability, nor the evidence base for use at the scale required. We demonstrate the performance, utility and validity of PRADA, as potential solution to address the inequalities in outcomes at the end of life.

## Introduction

Health services face mounting demand from aging populations with increasingly complex care needs, pressures that are exacerbated by fragmented care and its associated clinical, financial and sustainability risks [1–4]. With the National Health Service (NHS) in the UK said to be at an “existential brink”, addressing this is fundamental to NHS Long Term Plan ambitions [4]. The End-of-life (EOL) phase has high rates of urgent care use and deaths-in-hospital [5,6], with £22 billion spent in the last year of life, with 81% of this on hospital care [7]. The National Institute of Clinical Excellence reviewed EOL evidence [8], producing guidance [9] and standards [10] leading to statutory commissioning [11]. Registration to an EOL register was highlighted as fundamental to earlier, proactive and better coordinated care [8, 12,13] but less than a third of people who die are on an EOL register and little improvement has been made in the last decade [12,13]

The current pathway to EOL registration (EOLR) requires new, systematic, well governed, and fully commissioned approaches. The Gold Standards Framework Surprise Question is a prognostic tool for identifying those at the end of life asking, “Would you be surprised if this patient were to die within the next 12 months?”. The Surprise Question has notable limitations. Although it reflects clinicians’ intuitive assessments of end-of-life prognosis, its performance varies substantially according to baseline mortality risk within the population studied, the prediction interval, disease context, care setting, and the assessor’s previous experience. Moreover, it is typically deployed ad hoc at the point of care, which restricts its usefulness for scalable, systematic, and proactive risk stratification” [14].

Digital health care technology could improve EOL identification [15]. Current mortality prediction tools [16], developed in situational or disease-specific cohorts, are of uncertain validity, and are not automated [17–20]. Digitised tools often use complex comorbidity, clinical, laboratory, and activity data, and have not been widely adopted [21], whilst the NHS EARLY, a digital EOL risk stratification tool, has no published outcomes.

The Proactive Risk-Based and Data-Driven Assessment of Patients at the End of Life (PRADA) is an end-to-end digital health care system using integrated data, statistically validated risk variables, advanced analytics, combining robotic process and machine learning, risk stratifying whole populations, to identify those at higher mortality risk. It asks an “electronic Surprise Question” as a preliminary clinical decision support screen for clinicians. It triages those requiring further assessment for EOL care planning and EOLR and should thus drive better care. Evidencing the accuracy of advanced data methodologies [22], through tested and published validity, [23] is imperative. Prior PRADA outcomes confirm strong validity [24–29] but still required large-scale application to demonstrate performance validity and evidence of improved EOLR, unscheduled urgent care, and death-in-hospital rates. Our overall aim was to analyse the adverse consequences associated with missed EOLR and to evaluate performance of PRADA at scale in reducing inequalities in outcomes.

## Results

Of 12 practices, one was inactive but retained in analysis on an intention basis. In the baseline 2 years there were 64,887 individuals (alive 63,344 (97.6%), died 1,543 (2.4%)) and 66,321 (alive 65,496 (98.7%), died 831 (1.3%)) in the intervention year. The PRADA EOLR_BOT dashboard was used to risk escalate patients to clinicians for their assessment.

### Comparing those who died registered vs unregistered

**Table 1** shows outcomes in those who died. Over the whole 3 years, those registered were more likely to be older, female, have comorbidities, reside in a nursing home, have a higher frequency of recorded EOL processes and higher mortality probability estimations. Despite this, they had significantly lower rates of A/E attendance and non-elective admissions, reduced death-in hospital rates and 3- and 6-month hospital bed occupancy. Those not on the register still had high rates of recorded EOL processes and mortality probability estimates and were largely identified by the EOL-BOT (78.6%) and EOL-Tracker (96.1%) - the latter including those identified by the mortality probability prediction.

**Table 1.**
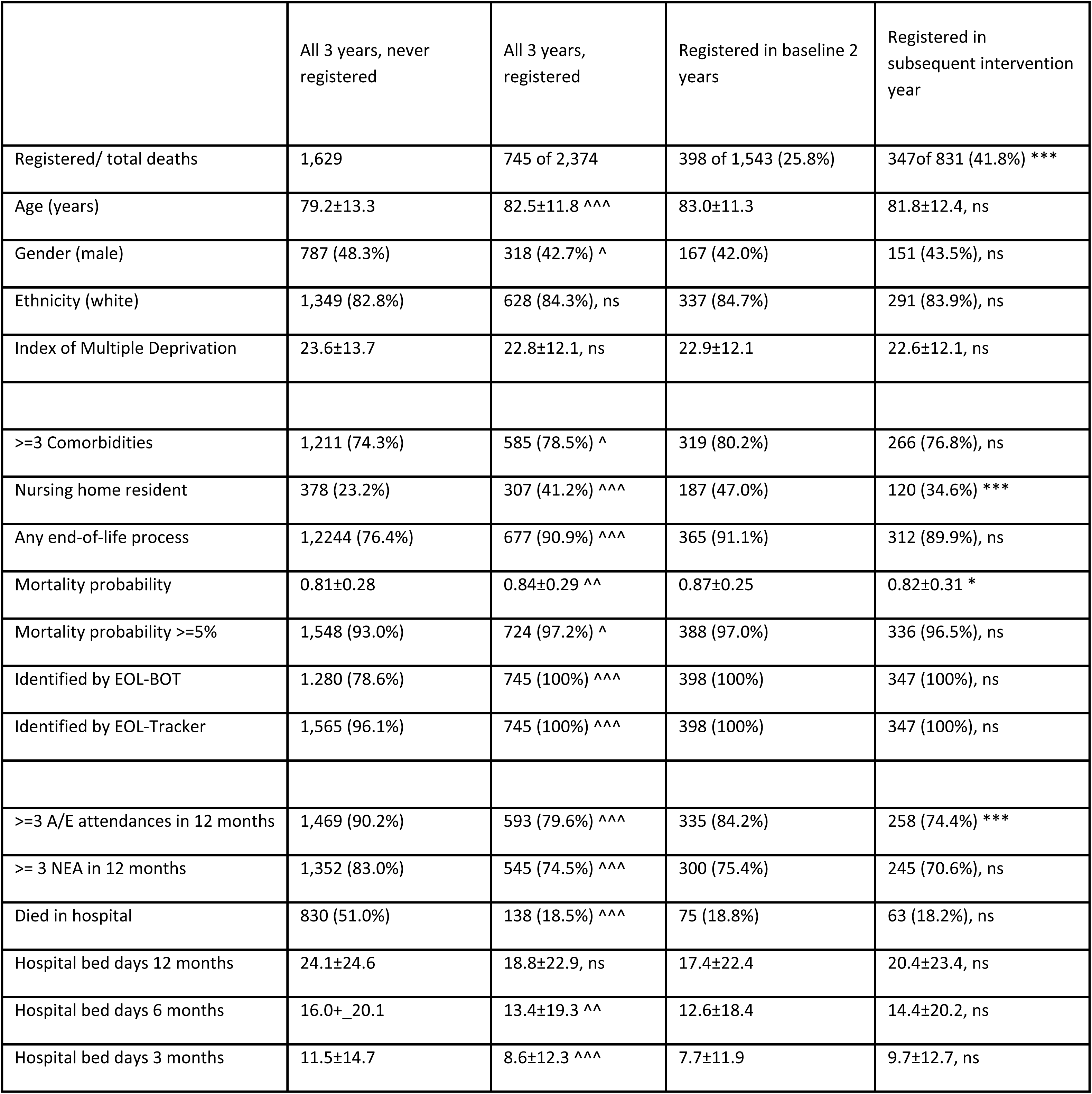
The outcomes amongst those who died by End-of Life registration status over 2 baseline years and a follow-on intervention year using the PRADA tool. Those never registered are compared to all who were registered over the whole 3 years (^). The baseline and post PRADA intervention cohorts of those registered are also compared (*). Data are the mean with its standard deviation or numbers with percentages. Statistical comparison is by the students t-test and the Chi-square tests for means and proportions, respectively. Single, double, and triple symbols = p<0.05, p<0.01 and p<0.001, ns = non-significant, EOL = end-of-life, A/E = accident and emergency department, NEA = non elective admissions.

|  | All 3 years, never registered | All 3 years, registered | Registered in baseline 2 years | Registered in subsequent intervention year |
| --- | --- | --- | --- | --- |
| Registered/ total deaths | 1,629 | 745 of 2,374 | 398 of 1,543 (25.8%) | 347 of 831 (41.8%) *** |
| Age (years) | 79.2±13.3 | 82.5±11.8 ^^^ | 83.0±11.3 | 81.8±12.4, ns |
| Gender (male) | 787 (48.3%) | 318 (42.7%) ^ | 167 (42.0%) | 151 (43.5%), ns |
| Ethnicity (white) | 1,349 (82.8%) | 628 (84.3%), ns | 337 (84.7%) | 291 (83.9%), ns |
| Index of Multiple Deprivation | 23.6±13.7 | 22.8±12.1, ns | 22.9±12.1 | 22.6±12.1, ns |
| >=3 Comorbidities | 1,211 (74.3%) | 585 (78.5%) ^ | 319 (80.2%) | 266 (76.8%), ns |
| Nursing home resident | 378 (23.2%) | 307 (41.2%) ^^^ | 187 (47.0%) | 120 (34.6%) *** |
| Any end-of-life process | 1,224 (76.4%) | 677 (90.9%) ^^^ | 365 (91.1%) | 312 (89.9%), ns |
| Mortality probability | 0.81±0.28 | 0.84±0.29 ^^ | 0.87±0.25 | 0.82±0.31 * |
| Mortality probability >=5% | 1,548 (93.0%) | 724 (97.2%) ^ | 388 (97.0%) | 336 (96.5%), ns |
| Identified by EOL-BOT | 1,280 (78.6%) | 745 (100%) ^^^ | 398 (100%) | 347 (100%), ns |
| Identified by EOL-Tracker | 1,565 (96.1%) | 745 (100%) ^^^ | 398 (100%) | 347 (100%), ns |
| >=3 A/E attendances in 12 months | 1,469 (90.2%) | 593 (79.6%) ^^^ | 335 (84.2%) | 258 (74.4%) *** |
| >= 3 NEA in 12 months | 1,352 (83.0%) | 545 (74.5%) ^^^ | 300 (75.4%) | 245 (70.6%), ns |
| Died in hospital | 830 (51.0%) | 138 (18.5%) ^^^ | 75 (18.8%) | 63 (18.2%), ns |
| Hospital bed days 12 months | 24.1±24.6 | 18.8±22.9, ns | 17.4±22.4 | 20.4±23.4, ns |
| Hospital bed days 6 months | 16.0+ _20.1 | 13.4±19.3 ^^ | 12.6±18.4 | 14.4±20.2, ns |
| Hospital bed days 3 months | 11.5±14.7 | 8.6±12.3 ^^^ | 7.7±11.9 | 9.7±12.7, ns |

### Comparing baseline to post PRADA use intervention

Comparing those who died whilst on the EOL register during the baseline 2 years to those who died during the intervention year, EOLR rates were 25.8% vs 41.8% (p<0.001) respectively (**Table 1**). The increase by overall practices in EOL registration was 17.6% ±13.7% (range -1.4% to 52.6 (3.4% to 52.6% in the 11 active practices)) (**Figure 1, top panel**). AE attendance was lower (74.4% vs 84.2%, p<0.001) whilst percentage rates of urgent admissions, death-in-hospital, and mean bed days occupied were similar. Increasing the proportion of patients on the EOLR from 25.8% to 48.1%, for the final year cohort of 831 patients, captured an additional 133 deaths, avoided 24 deaths-in-hospital, saved 386 bed days, with a cost avoidance of £192,789 at £500 per bed day. Had an ascertainment of 50% been achieved, hypothetically, an additional 68 would have been identified, avoiding a further 12 deaths-in-hospital, 198 bed days, and £98,806 costs.

**Figure 1.**
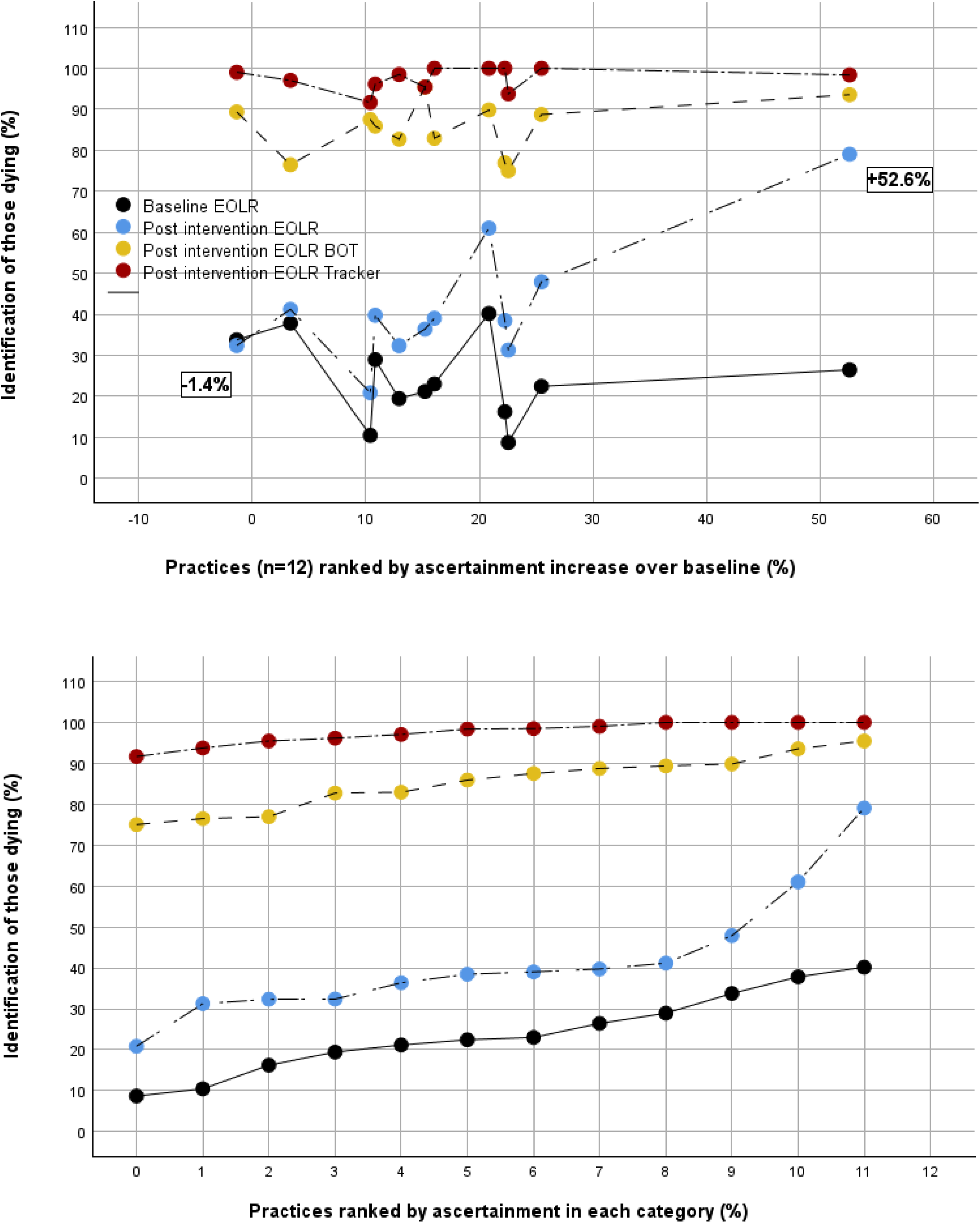
Shows the ascertainment of those who died over the baseline 2 years or the subsequent PRADA intervention year by actual end-of-life register (EOLR), or the 2 described methods used to predict EOLR. In the top panel, individual practice is ranked by improvement over baseline in the actual EOLR. In the bottom panel practices are ranked by degree of ascertainment in each of the 4 identification categories.

### Potential ascertainment gap

As seen in **Figure 1** (top panel), in the intervention year, the EOL-BOT identified 85.4+-6.7 (75.0 – 95.5) % of those dying by individual practice an increase over the formal EOLR of 43.7+-14.1 (14.5 – 66.7) %, (p<0.001). For the EOL-Tracker it was 97.5+-2.8 (91.7 – 100) %, increased 55.9+-14.1 (19.4-70.8) %, (p<0.001). The difference between EOL-BOT and Tracker was small but significant (12.1+-7.1 (0 – 23.1) %, p<0.001.

### Inter-practice variation

Taking percentage EOLR, EOLR-BOT and the EOL-Tracker in those dying as the dependent variable, practices, ranked by percentage ascertainment in each category, were entered into binary logistic regression with age, gender, ethnicity, deprivation score, comorbidity, and nursing home residence, but not variables influenced by EOLR (urgent care, care process completion). Using the lowest ranking practice as comparator, this produced demographic and case mix adjusted significance for the ascertainment variation by practice (**Figure 1**, bottom panel). For the baseline 2-year EOLR (*X^2^*=183.6, p<0.001), nursing home, practice and age were significant variables, explaining 11.6% of the variance, of which practices accounted for 31%. For the post-intervention EOLR (*X^2^*=96.6, p<0.001), nursing home and practice were significant, explaining 14.8% of variance, with practices accounting for 47.3%. For variation by the EOLR_BOT (*X^2^* 63.1, p<0.001), the model explained 13.6% of the variance, all due to demographic factors (age, comorbidity, nursing home residence) but not practice ranking. For the EOL-Tracker model (*X^2^*=5.4, p<0.05), the residual variance was minimal (4.3%) and was fully explained by age (p<0.05) but not any other demographic or case mix variable and not by between practice variation.

## Discussion

### Summary

Crucial to better end-of-life care is earlier identification for end-of life registration, but this must lead to better outcomes. In this study, in those that died prior to PRADA EOLR was low. Failed registration was associated with adverse outcomes. After PRADA implementation, EOLR improved, AE attendance was further reduced, and death-in-hospital rates and bed days occupied remained reduced, leading to cost savings. Independent of demographics and case mix, variation in EOLR was mostly explained by individual practice differences, even after improvement in EOLR . The use of two EOLR prediction methods adjusted for case mix and were not subject to inter-practice variation. A target of 50% EOLR in those that die is proposed as a realistic key performance indicator. This large-scale demonstration of improved EOLR outcomes in clinical practice strongly evidences PRADA’s utility and performance validity.

### Strengths and limitations

PRADA’s strengths are its cross-provider integrated dataset, analytical approach, risk stratification of whole populations, deployment in-real life primary care, function as a decision support aid and evidenced positive outcomes and validity. The study weakness is that it was not a randomised controlled trial with a parallel non-intervention limb.

### Comparison with existing literature

EOL prediction tools are mainly non-digital, non-scalable, or are disease specific and limited to hospital patients. There are comparable systems for EOL risk-stratification that could benchmark PRADA for criterion validity. The NHS commended EARLY programme has many similarities, but no published outcomes, whilst the John Hopkins Aggregated Diagnosis Groups (21) strongly predicted prospective whole population mortality (c-statistic 0.92) but there are no further publications since 2011 and real time function in front line clinical practice is unreported.

### General discussion

Although PRADA implementation resulted in notable improvement in EOLR there remains scope to optimise this system. Unregistered patients had high rates of co-morbidity, evidence of EOL process markers and high PRADA mortality probability estimates. Closing this gap in ascertainment by using the PRADA’s subsidiary identification tools, the EOLR-BOT or Tracker, could drive a shift from a reactive, late, one-to-one care model to proactive, early identification and one-to-many system. However, that has crucial dependencies.

### Clinician confidence

Clinical judgment is the mandatory gateway to EOLR in any EOL system. The conventional approaches to clinician engagement using information, education, and training-based quality improvement programs rely on the transfer of knowledge and translation into action. When translation fails, it is labelled as clinical inertia, overlooking the competing priorities and pressures in primary care. Addressing inertia by urging clinicians to “work harder” or expanding capacity to compensate for inefficiencies is an untenable approach. In contrast, service activation models are built around enablement. Such systems are structured to trigger action, requiring minimal additional knowledge. They initiate an action-driven cycle, where each act of care delivery builds experience, and positive outcomes. This shifts the burden from individual behaviour to system facilitated action, making pertinent interventions automatic and scalable, vital in high-pressure clinical environments.

Thus, variation in clinical decision making can be addressed, whilst allaying the anxieties of clinicians about more work of uncertain benefit under greater scrutiny. PRADA’s purpose is to promote efficiency through service activation, utility for clinicians, evidencing improved care, implemented with respect for clinical judgment.

So that clinicians might trust the system, there must be: a distinction between decision support and protocol enforcement; mitigation of any perceived threat to clinical autonomy and the patient’s wishes; and collaboration in system design. We have shown that integrating clinical decision making into PRADA enhances risk stratification (24) but, in pilot work, clinicians selected only 25% of those BOT identified for EOLR (29), and that the unselected remained of high 1 year mortality (29). Thus, to support clinicians in their decision making, unquestionably an iterative process, with many “grey case” issues, PRADA now re-escalates unselected at-risk patients, so assessments, through repetition, can be dynamic over altering illness trajectories. It also now incorporates an in-built critical event analysis tool so the system and its users can learn; and its reporting structures provide benchmarking against peers. Equally, for clinician buy-in, digital systems must have evidenced validity and we have shown PRADA to have face, content, concurrent, predictive and construct validity [24–29]. Performance validity with evidence of benefit in full operational deployment was limited [29] but now extends to 66,000 people manged in real-time primary care.

### Realistic ambitions and commissioning

Current guidelines recommended attainment of 1% EOLR in a whole population and 80% in those that die. The former is surely a reflection of the latter and will only show small shifts even if large gains are made in those that die. The 80% EOLR target in those that die, is of uncertain derivation, is unrealistic, being set beyond a likely maximal position. Accordingly, we have adopted a lower benchmark of 50% EOLR in those who die, confident that it can be attained or exceeded. In the 2 practices attaining >50% EOLR in those that died, in those currently alive, EOLR, EOL-tracker prevalence, and EOLR in the EOL-Tracker groups were 2.5% (mean), 4.4% and 56.6%. PRADA methodology thus offers realistic, peer comparable, case-mix adjusted, quantitative measures for governing and commissioning EOL register attainment.

### Enhanced support

Earlier, more accurate identification must lead to a direct, effective, one-to-one intervention. Beyond the digitally supported clinical decision, this will include complex and difficult conversations, the ascertainment of consent, the completion of relevant EOL processes including advance care planning and actual EOLR, and then regular review and ongoing support. Minimally doubling of EOLR rates thus presents a significant expansion of workload in primary and community care which needs quantifying, funding and commissioning.

### Implications for research and practice

Research communities must verify and enhance our findings with a particular need to understand workload and capacity requirements, the health economics of funding shifts to inform the realignment proposals of the NHS 10-year, plan, and the paramount need for qualitative research to engage patients to ensure respect of their autonomous wishes. Digital systems must engender clinicians’ confidence, but clinicians must drive implementation into practice, for which commissioners and policy makers must comprehend the wider transformational shifts required in care delivery and workload capacity to extract health economy wide efficiencies of manifest better care. Surely, strongly evidence based and validated digital healthcare methods must be rapidly and cohesively deployed at national scale, overcoming the vagaries, variations, and delays of local commissioning in the fragmented NHS, which will require leadership, ambition, direction, and cohesion at the highest level.

## Conclusion

In conclusion we confirm the extent of failed EOLR, its adverse outcomes, and that the variation in EOL_R rates links closely to service level variation. We demonstrate that PRADA’s support of clinical decision-making in front-line care, improves EOLR, with associated reductions in urgent care events. An inequitable care gap, which can be ascertained, continues to exist. Closing that gap has potential to reduce AE attendance, non-elective admission, death-in hospital, and hospital bed occupancy with financial savings. Variation in clinical decision making can be addressed positively using an activation approach respecting clinician autonomy. Realistic evidence-based benchmarks should be set for performance management, commissioning, and incentivisation, suggesting attained 50% EOLR in those that die as a new national target.

## Methods

### Study population

This large-scale, real-world, epidemiological cohort study was set in Wolverhampton, a deprived, multi-ethnic, industrial UK city. Twelve of 37 Wolverhampton practices participated in the programme. This 3-year study comprised 2 years before implementation of PRADA which included baseline EOL outcomes and 1 year post introduction of PRADA.

### Research questions

To determine in those who died: 1) the prevalence of failed end-of-life registration; 2) adverse outcome of non-registration; 3) PRADA’s ability to identify those at the end-of-life, the impact on EOLR rates, and ensuant urgent care outcomes.

### Data

The data set include the whole population aged >=18-years, registered to Wolverhampton general practices, including all deaths 3 years prior to analysis point without exclusion, sub-selection, or randomisation. Primary care, hospital, and community services data were linked under General Data Protection Regulation. The risk-stratification variables were demographics and those representing urgent care use, multimorbidity, care complexity and end-of-life markers: >=3 Accident and Emergency admissions over the prior 12 months; >= 3 non-elective admissions over the previous 12 months; >=3 co-morbidities, nursing home residency; markers of EOL needs included EOLR and any recorded EOL process (Electronic Palliative Care Co-ordination Systems (EPaCC), Recommended Summary Plan for Emergency Care and Treatment (ReSPECT), advanced care plans, contact with specialist palliative care teams and any known clinical assessment of EOL prognosis, captured in electronic patient records by the Surprise Question as “Would you be surprised if this patient died within the next year? = “No”. Demographic variables included age, gender, ethnicity, and the Index of Multiple Deprivation. The comorbidities utilized were the long-term conditions incorporated into UK primary care Quality Outcomes Framework. Variables were binary coded as 0 or 1 for absent vs present. No data were excluded or replaced by any assumed values. Mortality was determined from hospital mortality statistics and NHS Strategic Tracing Service checks.

### Statistical method

Data were analysed on IBM SPPS version 29. The student t-test and the Chi-square test were used for the difference between means and proportions, respectively. The association of independent variables with outcomes, and the production of mortality probability scores, was by binary logistic regression. Performance metrics were determined in receiver operating curve (ROC) analysis. Results are the mean ± standard deviation or number with percentage. Statistical significance was taken at p<0.05.

### Calculations for mortality probability, and theoretical EOL Registers

This methodology, including prediction of prospective mortality, is previously published [27,28]. It is repeated here in the current whole population 3-year data (264,012 cases (alive 256,631 (97.2%), died 7,381 (2.8%)). Binary logistic regression determined the association of vital status with age, gender, ethnicity, deprivation score, urgent care activity, comorbidity, nursing home residence, EOL registration and recorded EOL process (*X^2^* = 52,972.5, r^2^ = 0.81, p<0.001). It produced a mortality probability for everyone, whether alive or dead (0.00724+-0.05117 vs 0.75 +-0.32 respectively (p<0.001)). This was dichotomised at the 5th centile value of those who had died. This cut point identified 6,321 (2.5%) of those alive and 7,012 (95%) of those who died.

Two methods, descriptive and predictive, were used to create subsidiary cohorts of high mortality risk. The first, descriptive, termed the EOLR-BOT, used robotic process assessment (RPA), combining the known EOLR with any recorded EOL process, identifying individuals likely already on the EOL pathway, numbering 10,822 (4.1%) (alive 4,714 (1.8%), died 6,108 (82.8%)), of whom 4,773 (44.1%) were on the EOLR. The second, predictive, termed the EOL-Tracker, combined the RPA method with the dichotomised mortality probability score, identifying 15,736 (6.0%) (alive 8,567 (3.3%), died (7,169, 97.2%), EOLR 30.3%). The differences between the established EOLR, the EOL-BOT (6,049, 27% increase) and the EOL-Tracker (10,963 (129% increase) represent an ascertainment gap of those who required clinical assessment for EOLR inclusion. **Figure 2** shows receiver operating curve (ROC) for each measure’s association with mortality and **Table 2** details the metrics (all differences p<0.001). Sensitivity shifted from 37.3% to 81.9% and 97.1% for the actual EOLR, the EOL-BOT and the EOL-Tracker, respectively. By either EOLR derivation, assessment would be required of 3 patients for each dying. In this study, the EOLR_BOT was used to risk escalate patients to clinicians, since it had prior pilot evaluation (29), whist, at the time, the EOL-Tracker had not (28).

**Figure 2.**
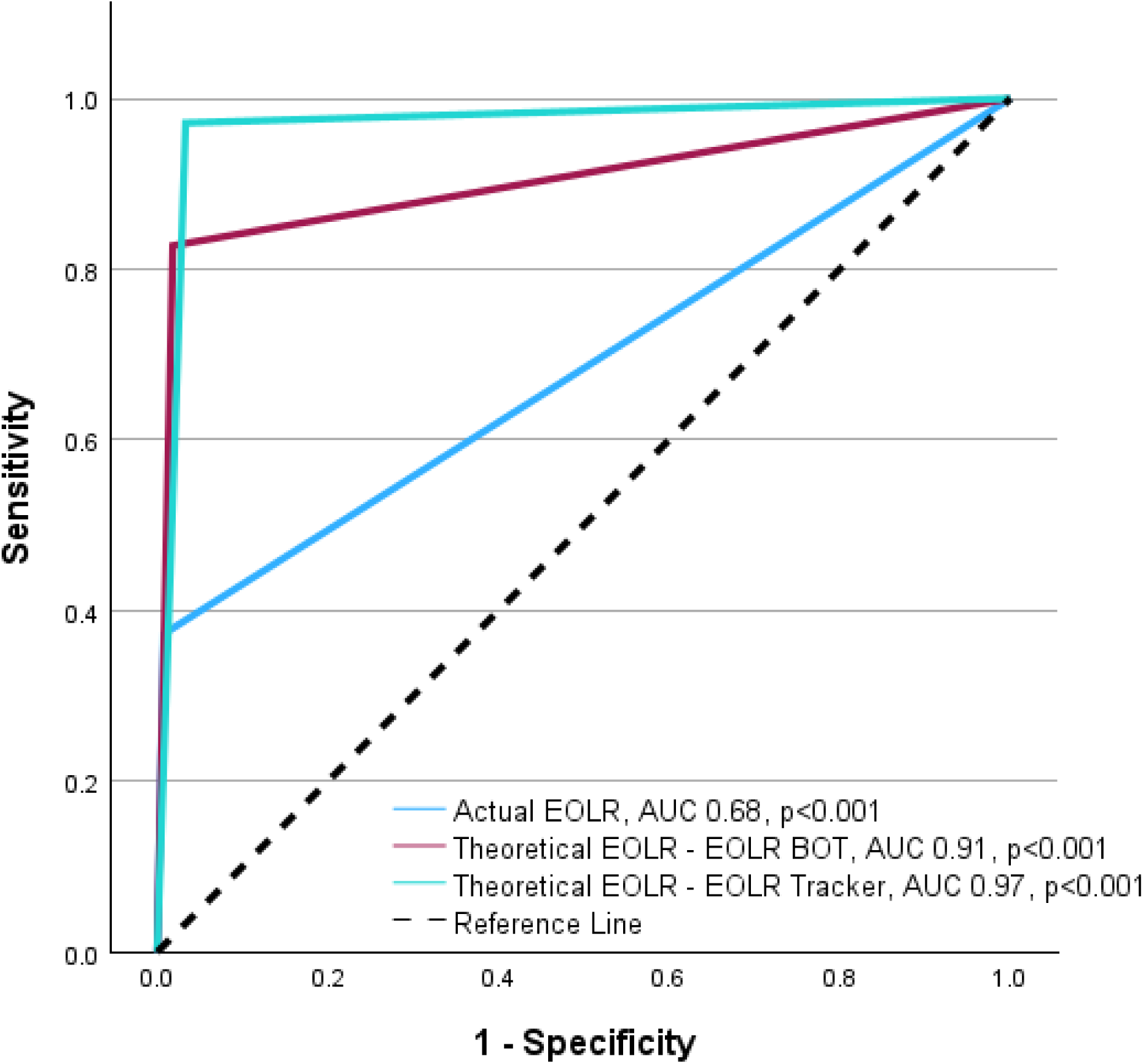
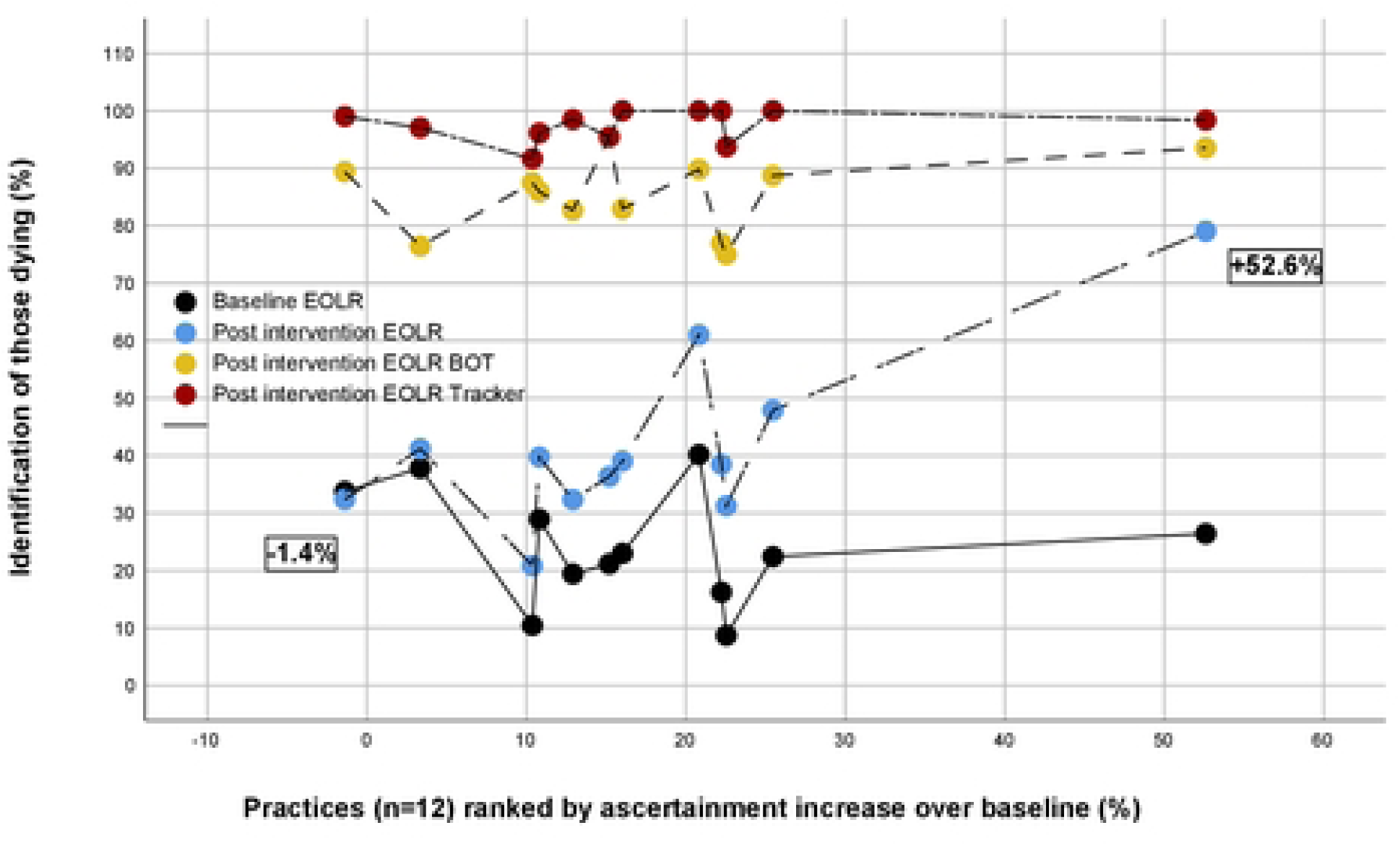
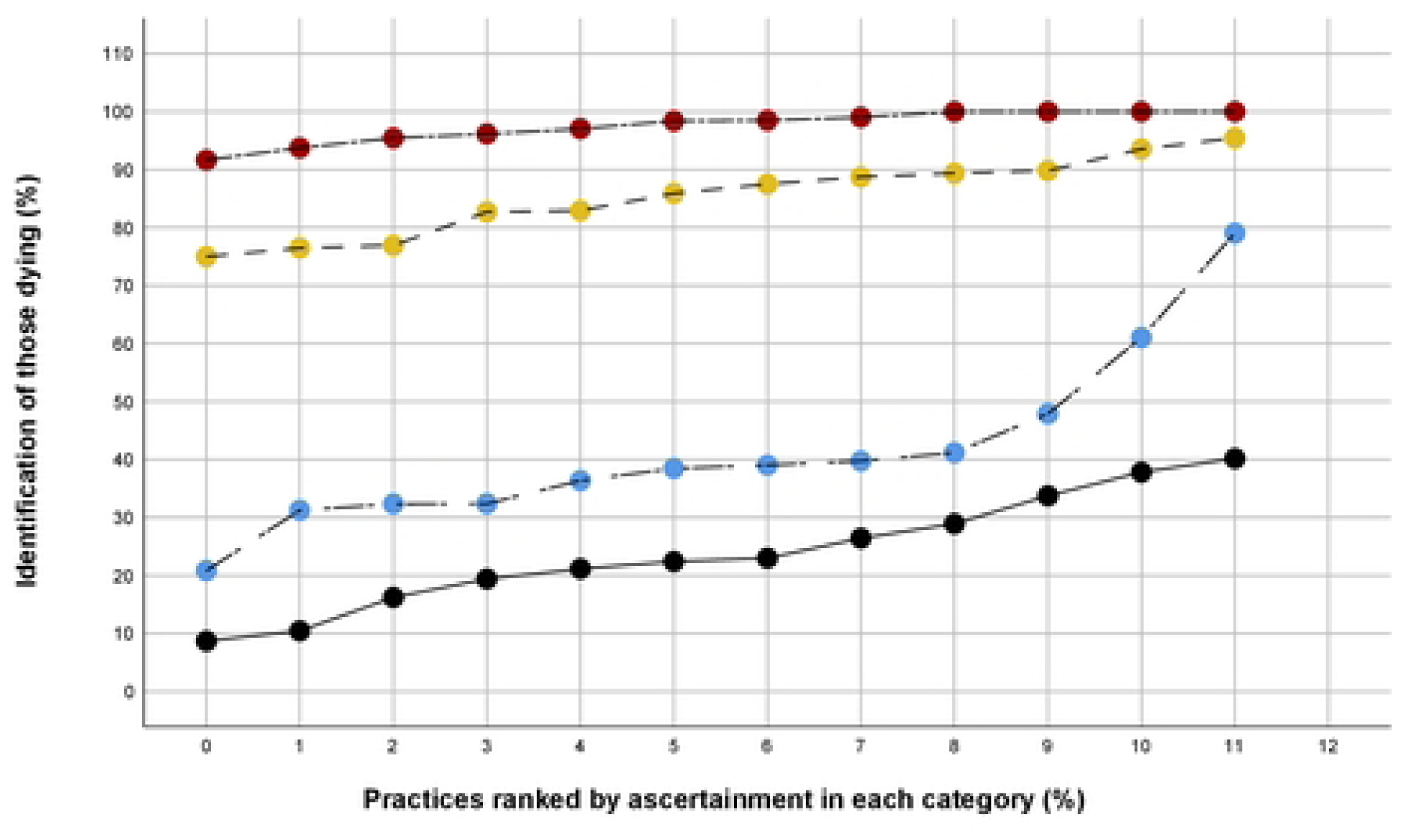
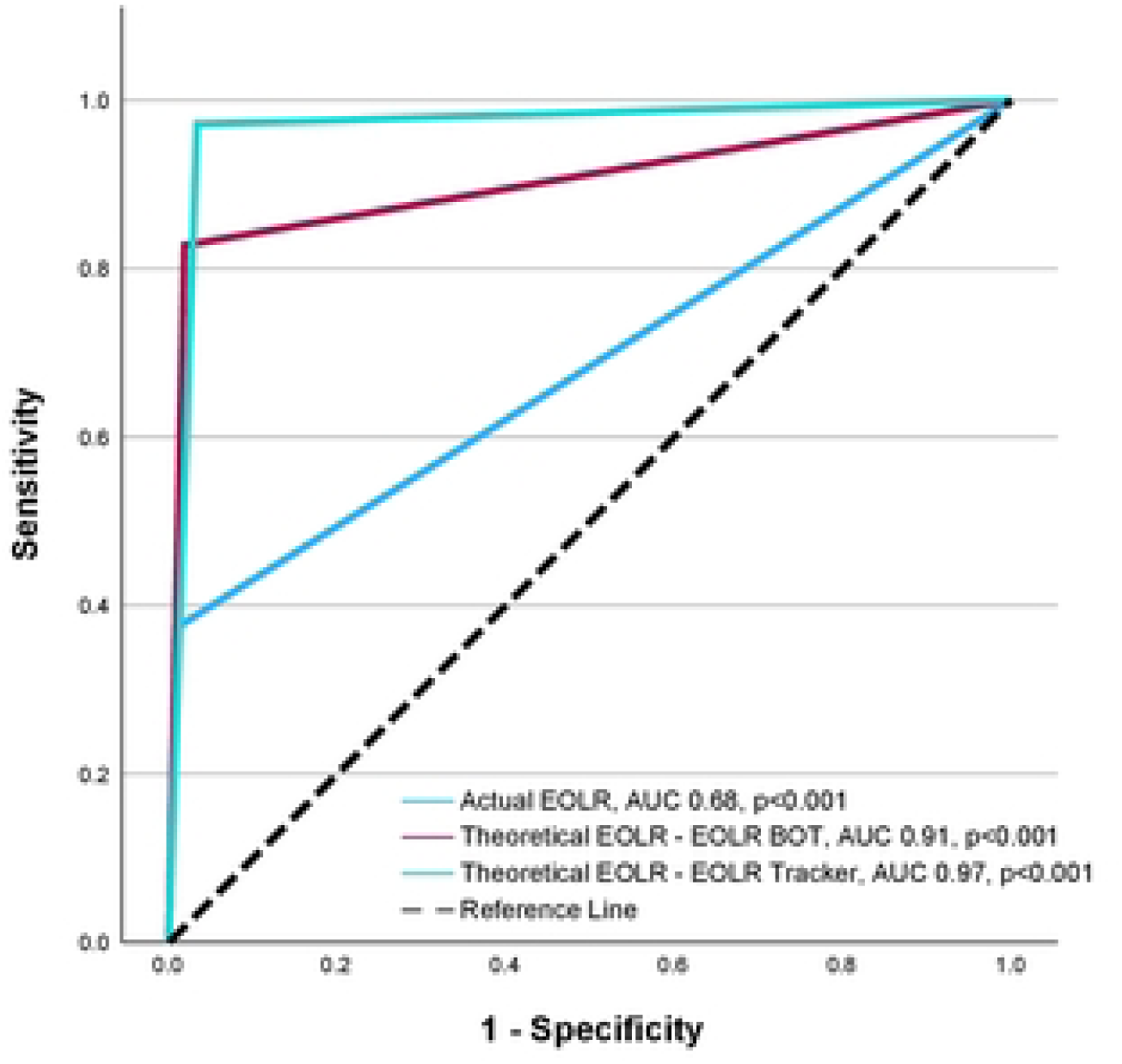
Receiver operating curves (ROC) for actual end-of-life registration (EOLR), and the 2 described methods used to predict EOLR and their association with mortality status in the whole 3-year data set (n= 264,012 adults (alive 256,631 (97.2%)). AUC = area under the curve, for which the ROC c-statistic is given.

**Table 2.** Receiver operating curve (ROC) performance metrics of actual end-of-life registration (EOLR), and the 2 described methods used to predict EOLR, and their association with mortality status in the whole 3-year data set (n= 264,012 adults (alive 256,631 (97.2%)). See Fig1.

|  | EOLR | EOLR-BOT | EOLR-Tracker |
| --- | --- | --- | --- |
| ROC c-statistic (95% CI) (all p<0.001) | 0.68 (0.67 - 0.69) | 0.91 (0.90 - 0.91) | 0.97 (0.967 -0.97) |
| Total identified (of 264,012) | 4,773 (1.8%) | 10,822 (4.1%) | 15,736 (6.0%) |
| True positive - deaths identified (of 7,381) | 2,751 (37.3%) | 6,108 (82.8%) | 7,169 (97.1%) |
| False positive | 2,022 | 2,022 | 8,567 |
| False negative - deaths missed | 4,630 (62.7%) | 1,273 (17.2%) | 212 (2.9%) |
| True negative | 254,609 | 251,917 | 248,064 |
| Sensitivity | 37.3% | 81.9% | 97.1% |
| Specificity | 99.2% | 98.1% | 96.7% |
| Positive predictive value | 57.7% | 56.5% | 45.6% |
| Negative predictive value | 98.2% | 99.5% | 99.9% |
| Accuracy | 97.5% | 97.7% | 96.7% |
| True positive: All identified | 1:1.7 | 1: 1.8 | 1: 2.2 |

### PRADA, the Proactive Risk-Based and Data-Driven Assessment of Patients at the End of Life, system

PRADA [24–29], built on integrated data and statistically significant variables, combines RPA with machine learning, “diagnosis vs prediction” or “what is vs what might be” [30], to risk stratify large scale populations [28]. Outcomes are presented in live direct care as a clinician decision support aid to promote “1 to many” anticipatory care. The clinical decision is recorded and used as a system variable [24]. Identified patients are then digitally followed for completion of EOL processes, including EOLR. Outcomes are ascertained prospectively.

### Research checklists

Strengthening the Reporting of Observational Studies.

### Ethical Considerations

This was a quality improvement initiative and did not involve randomisation or any intervention that ought not to have otherwise happened. Research ethical approval was not necessary, as confirmed by the authorised local research governance group.

### Patient consent for publication

Not applicable.

### Patient and Public Involvement

The use of clinical data in digital health care methodologies was discussed with and supported by the patient advisory and liaison group of our hospital.

## Data Availability

The corresponding author will share anonymized study data upon reasonable request, subject to approval of any proposal by the authors and our research governance body.

## Acknowledgements

The Wolverhampton Digital Heath Primary Care Research Network is a group of collaborating Wolverhampton GP practices implementing this programme in real time direct care, contributing to its development, clinical deployment, and continuous improvement, without which this ongoing innovation project would be impossible. The individual GPs are Drs I Adam, K Ahmed, J Burrell, AJ Cook, U Jaswal, A Koodaruth, C Madzima, G Malhi, G Pickavance, E Qureshi, M Sidhu. Contacts

## Competing or Conflicts of Interest

None

## Data Sharing

The corresponding author will share anonymised study data upon reasonable request, subject to approval of any proposal by the authors and our research governance body.

## Copyright

The corresponding author has the right to grant on behalf of all authors, the license to permit this article to be published.

## Abbreviations Used

EOL: End of life
EOLR: End of life register
NHS: National Health Service
PRADA: Proactive Risk-Based and Data-Driven Assessment of Patients at the End of Life
ROC: receiver operating curve
RPA: robotic process assessment

## Keywords

- Advance Care Planning (D032722)
- Digital Health (D000097103)
- End-Of-Life Care (D013727)
- General Practice (D058006)
- Health Inequities (D000091682)
- Health informatics (D008490)
- Hospital Mortality (D017052)
- Mortality (D009026)
- Palliative care (D010166)
- Quality Improvement (D058996)

## Author contributions (CRediT)

| Term | Definition | Authors |
| --- | --- | --- |
| Conceptualization | Ideas; formulation or evolution of overarching research goals and aims | BMS, VK, EP |
| Methodology | Development or design of methodology; creation of models | BMS, VK, EP |
| Software | Programming, software development; designing computer programs; implementation of the computer code and supporting algorithms; testing of existing code components | BMS, VK, WDHPCRN |
| Validation | Verification, whether as a part of the activity or separate, of the overall replication/ reproducibility of results/experiments and other research outputs | BMS, AT, EH, EP |
| Formal analysis | Application of statistical, mathematical, computational, or other formal techniques to analyze or synthesize study data | BMS, |
| Investigation | Conducting a research and investigation process, specifically performing the experiments, or data/evidence collection | WDHPCRN, BMS, JL, VK |
| Resources | Provision of study materials, reagents, materials, patients, laboratory samples, animals, instrumentation, computing resources, or other analysis tools | N/A |
| Data Curation | Management activities to annotate (produce metadata), scrub data and maintain research data (including software code, where it is necessary for interpreting the data itself) for initial use and later reuse | BMS, VK |
| Writing - Original Draft | Preparation, creation and/or presentation of the published work, specifically writing the initial draft (including substantive translation) | BMS, EP, AT, EH |
| Writing - Review & Editing | Preparation, creation and/or presentation of the published work by those from the original research group, specifically critical review, commentary or revision – including pre- or postpublication stages | All |
| Visualization | Preparation, creation and/or presentation of the published work, specifically visualization/ data presentation | BMS |
| Supervision | Oversight and leadership responsibility for the research activity planning and execution, including mentorship external to the core team | BMS, EP |
| Project administration | Management and coordination responsibility for the research activity planning and execution | JL |
| Funding acquisition | Acquisition of the financial support for the project leading to this publication | BMS |

